# Long-term Associations of COVID-19 Severity with Cognition and Mental Health in an Indian Cohort

**DOI:** 10.1101/2025.11.17.25340384

**Authors:** Aruneek Biswas, Vivek Pamnani, Priyanka Srivastava, Vishnu Sreekumar

**Affiliations:** Cognitive Science Lab, International Institute of Information Technology, Hyderabad, India

**Keywords:** COVID-19, Indian population, COVID-recovered, long-term associations, cognition, mental health

## Abstract

Long-term cognitive outcomes following COVID-19 remain poorly characterized in South Asian populations. We assessed 181 Indian adults—99 with a reported history of COVID-19 at least six months earlier and 82 healthy controls—using standardized tests of working memory, selective attention, and pattern separation, alongside validated mental-health scales. Disease severity was assessed as per WHO guidelines rather than by degree-of-hospitalization, capturing individuals who experienced severe illness outside hospital care. Those with moderate-to-severe COVID-19 showed pronounced working-memory deficits compared with mildly infected and uninfected controls, while selective attention and pattern separation were comparable across groups. Vaccination at the time of infection was associated with better pattern separation, suggesting selective hippocampal protection. Notably, nearly all participants with moderate-to-severe illness reported a complete recovery despite continuing to exhibit measurable cognitive deficits. However, we did not observe any robust differences in affect or in the propensity toward depression and anxiety. Taken together, our findings suggest that hospitalization is an inadequate proxy for disease severity and that subjective recovery is not an appropriate yardstick for assessing Long COVID, underscoring the need for context-sensitive, population-based assessments of post-COVID sequelae.

## 1. Introduction

The COVID-19 pandemic has precipitated a global health crisis, with the long-term sequelae of SARS-CoV-2 infection emerging as a significant public health challenge. Beyond the acute respiratory illness, a substantial number of individuals experience protracted symptoms, a condition commonly referred to as “Long COVID” (Greenhalgh et al., 2024). Among the most persistent and debilitating of these symptoms are the neurocognitive and psychiatric manifestations (Natarajan et al., 2023; Aretouli et al., 2025). Patients frequently report “brain fog,” characterized by difficulties with attention, concentration, and memory, which can profoundly impact daily functioning and quality of life (Aretouli et al., 2025; Jain et al., 2023).

A large-scale study by Taquet et al. (2022) assessed neurological and psychiatric risks after COVID-19 of over a million patients from their health records by propensity-matching them to controls with respiratory infections and identified significantly higher risks of cognitive deficit in adults, even at two years post-infection. Likewise, Elboraay et al. (2025), in a recent meta-analysis, reported that approximately 27.1% of patients exhibited cognitive impairment and 27.8% exhibited memory disorders six months or more after infection. Latifi and Flegr (2023) found that young adults recovering from severe COVID-19 exhibited fatigue, cognitive slowing, and deteriorated mental health months after infection. Similarly, Diana et al. (2023) provide longitudinal evidence of cognitive and psychological alterations in COVID-19 up to a year after infection.

Although considerable research has been dedicated toward understanding these impairments, the majority of studies have been focused on European and American populations, leaving a critical gap in the understanding of the long-term impact on the vast and diverse populations of the Global South (Fig. 1B in Hou et al. (2025)). This context is particularly important for developing tailored interventions and healthcare policies, as their unique demographic, genetic (Niemi et al., 2022), and socioeconomic factors (Jassat et al., 2023) may modulate the disease’s long-term trajectory. Therefore, a focused investigation into the long-term cognitive and mental health outcomes following COVID-19 within S. Asian populations is imperative.

**Figure 1:**
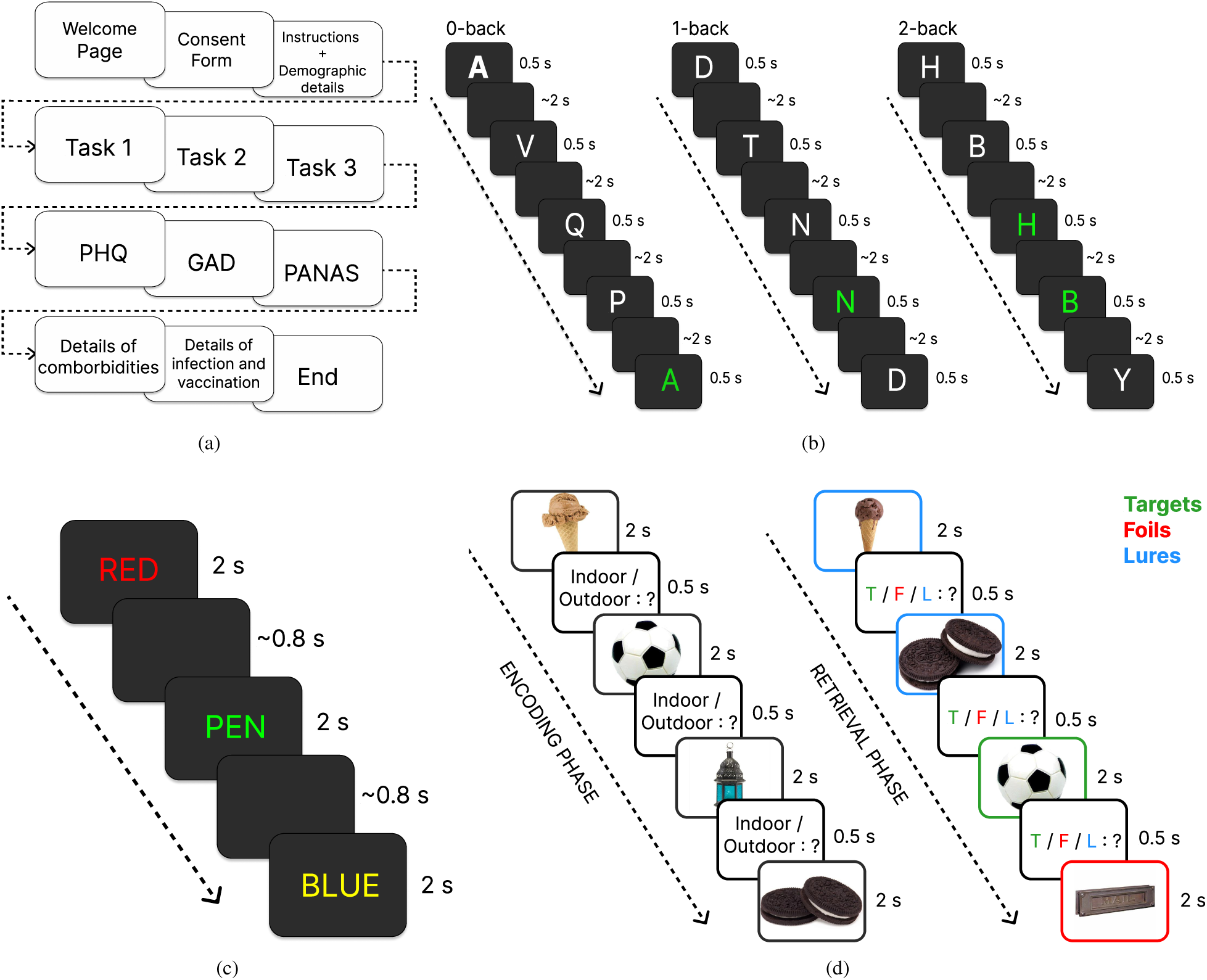
Overview of the experimental design and the three cognitive tasks. **(a)** Sequence of the administered battery. **(b)** n-back task schematic illustrating the 0-back, 1-back, and 2-back conditions. Letters highlighted in green indicate trials where participants were expected to press a key; this color-coding is for illustration only and was not present during the actual task. **(c)** Modified Stroop Task showing example stimuli from the congruent, neutral, and incongruent conditions. **(d)** Mnemonic Similarity Task (MST) showing the encoding and subsequent retrieval phases. Colored box boundaries denote stimulus types (targets, lures, foils) for illustration purposes only and were not visible to participants during the task.

To date, only a handful of studies have examined the long-term cognitive and mental health sequelae of COVID-19 in Indian or broader South Asian cohorts, and their findings remain inconsistent. Chakrabarty et al. (2024) reported substantial rates of depression, anxiety, and insomnia among hospitalized COVID-19 survivors in Kolkata, a year after their initial infection, alongside measurable cognitive deficits on the ACE-III and Trail Making Test, which were strongly correlated with psychiatric symptoms. In contrast, Gopinath et al. (2025) found minimal long-term impairments—limited to subtle inefficiencies in working memory and executive control—in a young adult cohort who had not required hospitalization for their infection and were tested nearly two and a half years later. Crucially, however, both studies relied on hospitalization status as a proxy for disease severity, a major limitation in the Indian context, where, during peak pandemic waves, many clinically severe cases went unhospitalized due to the overwhelming strain on the healthcare system.

Thus, in this study, we aimed to investigate how COVID-19 severity relates to long-term outcomes in working memory, pattern separation, and attentional control, assessed at least six months after infection using a clinically validated severity measure that ran independent of hospitalization. Given emerging physiological evidence that SARS-CoV-2 affects hippocampal circuitry (Nouraeinejad, 2023), we were especially interested in whether pattern separation, a process tied to hippocampal integrity, showed any significant alteration, a question that had not yet been examined as of the time of designing the study. Accordingly, we employed a battery of relevant cognitive tasks alongside mental health questionnaires measuring anxiety, depression, and affect to obtain a comprehensive assessment of cognitive function and mental well-being.

Given that our comparison involved subtler gradations of disease severity (rather than a stark hospitalized *vs.* non-hospitalized contrast) and we do not selectively recruit only those who report ongoing symptoms as of the assessment (that is, we do not use the definition of long-COVID as our inclusion criterion), we were uncertain whether clear severity-dependent cognitive deficits would emerge. Nevertheless, we expected that patients would show impairment proportional to the severity of the acute infection in some or all of the cognitive domains and mental health measures. Further, while the efficacy of vaccines in preventing severe COVID-19 is well established (Yang et al., 2023; Watanabe et al., 2023; Maier et al., 2024), their role in mitigating long-term cognitive and mental health sequelae (independent of their effect on disease severity) remains largely unexplored. In particular, few studies have used objective measures to assess these outcomes, contributing to this gap (Zhao et al., 2023). Therefore, we also sought to examine whether vaccination status at the time of infection was associated with long-term cognitive performance.

## 2. Method

### 2.1. Materials

#### 2.1.1. Overall Assessment

Participants completed an unsupervised computerized cognitive and mental health assessment battery using an experiment designed in PsyToolkit (Fig. 1a), either on-site at XXX premises or remotely via PsyToolkit Cloud using a non-mobile device (Stoet, 2010, 2017). At the outset, participants were instructed to complete the assessment alone, in a quiet room, and without distractions, including those from other digital devices. The data collection commenced on October 4, 2022 and concluded on October 6, 2023. It is prudent to note that internet connectivity issues do not affect response times, as PsyToolkit preloads the full experiment in the participant’s browser and records all timing locally; further, PsyToolkit experiments are lightweight and designed to run smoothly even on relatively old systems, reducing concerns arising from variation in computing hardware.

Socio-demographic details were collected at the outset (see Appendix A.1 for the questions). Participants completed three cognitive tasks — the n-back Task (*n* = 0, 1, 2) (Miller et al., 2009; Frost et al., 2021), Stroop Task (MacLeod, 1992; Algom et al., 2021), and Mnemonic Similarity Task (MST; (Stark et al., 2019)) — designed to assess working memory, selective attention, and pattern separation, respectively. Practice rounds were administered for all the tasks. Following this, they completed standard mental health assessments — Patient Health Questionnaire (PHQ; (Kroenke et al., 2003)), Generalized Anxiety Disorder (GAD; (Kroenke et al., 2010)), and Positive and Negative Affect Schedule (PANAS; (Watson et al., 1988)) — which measure mood, anxiety, and affect, respectively. Additionally, participants provided information about their personal and familial history of mental health disorders, details of any COVID-19 infections (including timing, severity, hospitalization status, and recovery duration), physical comorbidities, and their vaccination status at the time of the survey and their first infection (see Appendix A.2 for the questions).

#### 2.1.2. Cognitive Tests and Performance Metrics

##### (i) Working memory (n-back task)

In this study, we implemented three variations of the delayed-match-to-sample n-back task (NB-DMS; Fig. 1b): a 0-back condition (*n* = 0), as well as 1-back and 2-back conditions (*n* = 1, 2). Two blocks for each variant was administered and the order of the blocks was chosen at random. Each block comprised a sequence of 21 letters and there was a 33% probability that the current stimulus matched the one *n* trials back. Each letter was presented for 0.5 s, followed by a jittered inter-trial interval (ITI) ranging between 1.9 and 2.1 s; the response window extended across both the presentation window and ITI.

The 0-back task operated as a go/no-go paradigm, requiring participants to press a button whenever a predetermined target letter, specified at the beginning of the block, appeared on the screen. This condition serves to measure sustained attention (Miller et al., 2009). In contrast, the 1-back and 2-back conditions required participants to respond when the currently displayed letter matched the one presented one or two trials earlier, respectively; they serve to measure unstructured working memory span (Frost et al., 2021).

For all the variants, we computed *d^′^* (theoretical range: (*−*∞, ∞)) to assess task performance (Haatveit et al., 2010). It quantifies sensitivity by measuring the normalized distance between the probability distributions of hits and false alarms. The higher the metric, the better the performance.

##### (ii) Selective Attention (Serial Stroop Color-Word Interference Test)

There exists no consensus on the design of a digitally administrable Stroop task (Brunetti et al., 2021); so, we roughly reproduced Exp. 1B in (Salo et al., 2001) but switched the response modality from vocal to manual, for easier administration. This task serves as a reliable measure of selective attention but in light of recent evidence (Algom et al., 2021), we do not claim the task to be a good measure of conflict monitoring.

Our task consisted of 120 trials, divided equally under three conditions (Fig. 1c): (i) a color word being displayed in its corresponding ink color (congruent condition), (ii) a non-color word being displayed in one of the ink colors (neutral condition), and (iii) a color word being displayed in a non-matching ink color (incongruent condition). The trials were randomly intermixed and administered serially. For all trials, participants had to identify the ink color by pressing one of four buttons corresponding to Red, Green, Blue, or Yellow. Each stimulus was displayed for 2 s, during which participants could respond. This was followed by a jittered delay (0.45–0.55 s) to avoid entrainment effects, a fixation cross (0.2 s), and a blank screen (0.1 s) before presentation of the next stimulus, resulting in an intertrial interval of 0.75–0.85 s.

To assess task performance, we computed the percentage increase in reaction time (RTIE; theoretical range: (*−*100, ∞)) for attempted incongruent trials compared to attempted congruent trials, where lower values indicate better inhibitory control. Notably, the serial version of the task produces a smaller magnitude of interference effect than the manually administered blocked variant (Penner et al., 2012; Brunetti et al., 2021); further, the modality of manual response has been shown to reduce the interference effect compared to the classical vocal modality (Augustinova et al., 2019).

##### (iii) Pattern Separation (Mnemonic Similarity Task)

This task (Fig. 1d) involved two phases: the encoding phase and the testing phase. In the encoding phase, participants classified 80 stimuli as indoor or outdoor objects. In the retrieval phase, 120 stimuli were presented in random order, 40 being exact repetitions of images displayed in the previous phase (targets), 40 being new images not previously seen (foils), and the remaining being perceptually similar to those seen during the study phase, but not identical (lures; classified into five levels of increasing non-similarity from L1 to L5). Participants had to classify each stimulus as “old,” “similar,” or “new.” The task is considered a reliable assay of hippocampal functioning, particularly dentate gyrus-dependent fine-grained pattern separation (Stark et al., 2019).

In the encoding phase, each image was shown for up to 2 s, followed by a 0.5 s gap for the participant to make a classification choice; if participants responded early, a blank screen was displayed for the remaining time. Likewise, in the retrieval phase, each stimulus was displayed for 2 s, followed by a 0.5 s gap for the participant to log their response.

As a metric of task performance, we computed the Lure Discrimination Index (LDI; theoretical range: [*−*1, 1]), which is the difference between the probability of providing a “similar” response to the lures and the probability of providing a “similar” response to the foils (Stark et al., 2019). The higher the metric, the better the performance.

#### 2.1.3. Mental health surveys and metrics

i. The **Patient Health Questionnaire (PHQ)** is a self-report Likert-type questionnaire to assess the severity of depression symptoms (Kroenke et al., 2001). We initially administered the PHQ-2 (items 1 and 2 of the PHQ-9; score range: 0–6) to approximately half of the participants, later switching to the more comprehensive PHQ-9 (9 items; score-range: 0-27). Following standard practice, items were presented in their standard order without randomization. To ensure comparability across participants, scores were evaluated only for the two items common to both instruments (Kroenke et al., 2003). Higher scores reflect greater severity and likelihood of depression; a score of 3 or more on the PHQ-2 can be taken as a clinical cutoff for identifying individuals at risk of major depressive disorder (Manea et al., 2016; Levis et al., 2020).
ii. The **Generalized Anxiety Disorder (GAD)** is a self-report Likert-type questionnaire assessing anxiety symptoms over the past two weeks, derived from the DSM-IV criterion (Spitzer et al., 2006). We initially administered the GAD-2 (items 1 and 2 of the GAD-7; score range: 0–6) to approximately half of the participants, later switching to the GAD-7 (7 items; score range: 0–21) for a more comprehensive assessment. Following standard practice, items were presented in their standard order without randomization. To ensure comparability across participants, scores were evaluated only for the two-questions common to both instruments. Higher scores indicate greater severity of anxiety symptoms; a score of 3 or more on the GAD-2 is suggested as a clinical cutoff for identifying individuals at risk of generalized anxiety disorder (Plummer et al., 2016).
iii. The **Positive and Negative Affect Schedule (PANAS)** is a self-report scale test that contains ten questions and is used to assess positive and negative affect (Watson et al., 1988). Low PA is strongly linked to depression; high NA is weakly linked to both depression and anxiety (Watson and Levin-Aspenson, 2017).

### 2.2. Participants

Any Indian citizen, aged 18 to 60 years, was eligible for enrollment in our study. We did not explicitly target COVID patients to avoid self-selection bias. Color-blind subjects were screened out via the Ishihara test (Ishihara, 1917).

Participants were recruited through social media, email, participant referrals (snowball sampling), and the participant-recruitment platform CloudResearch^2^(Hartman et al., 2023). Snowball sampling involved participants circulating the study invitation within their personal, student, or institutional networks. Participants recruited through CloudResearch were accepted only if from India and compensated with USD 5, while those recruited through other sources received INR 200 for their time. This study was approved by the XXX Institute Review Board (IRB), and participants provided informed consent through the study website before beginning the assessment. A total of 256 participants were recruited across these sources.

Owing to the complexity of our statistical models, a priori sample size estimation for a predetermined effect size was challenging; while approaches exist for predictive modelling (Riley et al., 2018), these were overly conservative for our inferential objectives. We therefore adopted a pragmatic approach, informed by prior studies conducted in low-resource settings with comparable designs (Abdelghani et al., 2022; Ollila et al., 2022), and targeted a sample size of approximately 200 participants. This decision was further shaped by practical constraints, particularly the substantial difficulty in recruiting individuals with moderate and severe infection despite sustained recruitment efforts.

#### 2.2.1. Groups

The sample was broadly categorized into four groups. One group consisted of healthy controls who reported neither symptoms suggestive of COVID-19 nor a previous positive SARS-CoV-2 test; however, we concede that asymptomatic infections in this group could not be excluded to certainty. The other three groups included participants with a history of COVID-19 symptoms, ranked by the severity of those symptoms (reproduced verbatim from our survey; (COVID-19 Treatment Guidelines Panel, 2019)):

i. **Mild Illness:** Individuals who exhibit various signs and symptoms of COVID-19 (e.g., fever, cough, sore throat, malaise—a general feeling of discomfort, illness, or uneasiness—headache, muscle pain, nausea, vomiting, diarrhea, loss of taste and smell) but do not experience shortness of breath, dyspnea, or abnormal chest imaging.
ii. **Moderate Illness:** Individuals who show evidence of lower respiratory disease during clinical assessment or imaging and have an oxygen saturation (SpO_2_) *≥* 94% of room air at sea level.
iii. **Severe Illness:** Individuals with (SpO_2_) *<* 94% on room air at sea level, a ratio of arterial partial pressure of oxygen to fraction of inspired oxygen (PaO_2_/FiO_2_) *<* 300 mm Hg, a respiratory rate *>* 30 breaths/min, or lung infiltrates *>* 50%.

In total, 256 participants had completed the survey, but 55 participants had to be excluded for failing quality control checks and/or giving inconsistent data and/or being unqualified for our assessments, as elaborated in Appendix B. We merged the moderate and severe categories into one group (**mdrt+** henceforth) due to the extremely low number of severely infected participants; eleven participants who reported a positive diagnostics test but no COVID-19 symptoms were classified as confirmed asymptomatic infections and excluded from further analyses because of the low group size (Table 1).

**Table 1:** Medical severity profile of participants.

| Infection Severity | Healthy | Asymp | Mild | Mdrt | Svr |
| --- | --- | --- | --- | --- | --- |
| Total Count | 82 | 11 | 85 | 19 | 4 |
| Home Quarantine | 0 | 5 | 62 | 10 | 2 |
| Home Stay with Medical Care / Support | 0 | 5 | 22 | 7 | 0 |
| Hospitalization (General Ward / Special Ward) | 0 | 0 | 0 | 1 | 0 |
| Hospitalization (Intensive Care) | 0 | 0 | 0 | 1 | 0 |
| Continuous Oxygen supplement | 0 | 0 | 0 | 0 | 0 |
| Oxygen Support (not continuous) | 0 | 0 | 0 | 0 | 1 |
| Ventilator support | 0 | 1 | 0 | 0 | 1 |
| High-flow nasal support | 0 | 0 | 0 | 0 | 0 |

Further, because our analysis focused on the long-term effects of COVID-19, we excluded from all analyses six participants whose first reported infection occurred 26 weeks or less before the survey and three who did not report the probable timing of their infection. Four additional participants reported implausible first-infection dates preceding February 2020. However, ancillary infection history and vaccination information indicated that they had been infected more than 26 weeks before the survey. They were therefore retained in comparisons between infected participants and healthy controls but excluded from within-infected-group analyses requiring reliable infection timing. Thus, the former analyses included 181 participants (82 healthy, 76 mild, and 23 mdrt+), whereas the latter included 95 infected participants (75 mild and 20 mdrt+).

These severity criteria were developed for clinical use but were applied here through retrospective self-report rather than adjudication by clinician(s). The mild/moderate+ distinction was anchored by the reported absence versus presence of shortness of breath, a symptom that can be readily identified without any clinical expertise. Moderate and severe cases were pooled, eliminating the consequences of uncertainty in distinguishing between these two categories. We additionally compared reported severity with treatment course and excluded seven participants whose combinations were clearly inconsistent ( Appendix B). Nevertheless, some classification error remains possible.

### 2.3. Statistical Methods

The data was pre-processed in Python using Statsmodels (Seabold and Perktold, 2010) and SciKit (Pedregosa et al., 2011), and visualized using PtitPrince (Allen et al., 2021). Regression modeling and diagnostics were done exclusively in R using default functions, and the following packages: Ordinal (Christensen, 2023), boot (Angelo Canty and B. D. Ripley, 2024), and gofcat (Ugba, 2022).

Participant data for a cognitive task were excluded if they exhibited excessive non-responses in the task (>50% of trials; except in the n-back task where non-response can be a valid response) or unusually slow mean response times (*z*-score *>* 3), which likely reflected impaired task engagement or lack of diligence. To evaluate whether performance was above chance, we conducted a one-sided Wilcoxon signed-rank test (Q–Q plots revealed substantial deviations from normality in almost all cases), with the null hypothesis that the median of each metric distribution was equal to zero, tested at the 5% significance level.

We first derived composite cognitive and mental-health scores using PCA, and then compared cognitive and mental-health outcomes across COVID-19 severity groups using regression models appropriate to each outcome. Analyses contrasted infected participants with healthy controls and, separately, moderate-to-severe with mild cases, with prespecified covariate adjustment. Statistically significant moderate-to-severe effects were additionally subjected to sensitivity analyses.

#### 2.3.1. Composite Scores

To assess global cognitive performance across the tested domains, as in Hampshire et al. (2021), we extracted the Cognitive Composite Score (CCS) by applying Principal Component Analysis (PCA) on the standardized (z-scored) cognitive metrics, using Singular Value Decomposition, and taking the first component (explained *≈* 43% of the variance):

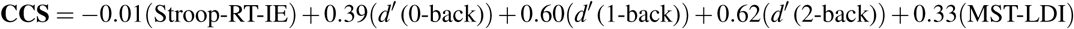

Cognitive Composite Scores were calculated for only those participants who attempted *all* five tasks. A higher CCS score corresponded to better cognition. Likewise, we extracted the Mental Composite Score (MCS) from the standardized (z-scored) mental health metrics described above

(explained *≈* 60% of the variance):

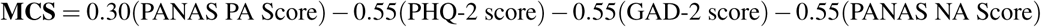

A higher MCS score signifies better mental health.

#### 2.3.2. Regression Modelling

i. **Cognitive Metrics:** To assess associations between COVID-19 severity and our individual cognitive outcomes as well as the Cognitive Composite Score (CCS), we modeled the metrics as follows:

a. *d^′^* **(n-back Task), RT-IE (Stroop Task), and Cognitive Composite Score:** Being unbounded and continuous, these metrics were modeled using Multiple Linear Regression Models (MLRM).
b. **LDI (MST):** We applied a linear transformation (*y^′^* = *<u>^y^</u>*<u>^+1^</u>) to map the outcome variables onto the interval (0, 1) and ran a Beta Regression Model (BRM) (Geissinger et al., 2022) with a logit link; no boundary values were observed in the original data and hence we didn’t require a zero/one inflated model. Numerous alternatives exist for modeling continuous bounded outcomes (Korkmaz, 2019), most are seldom used, lack mature software implementations, and require a level of theoretical grounding beyond the present scope; hence, we adopted the well-established Beta regression framework. We applied bias correction (Grün et al., 2012) to obtain more reliable estimates of the regression parameters. Two sets of comparisons were conducted:

a. Infected participants vs. healthy controls
b. Mdrt+ vs. mild cases, with additional control for time since first infection, number of infections, and vaccination status at the time of the first infection. This also enabled us to evaluate whether vaccination status was associated with cognitive performance after adjustment for disease severity and whether cognitive performance improved with time since infection. We included age, gender^3^, education, region, presence of physical comorbidities, and GAD and PHQ scores (as proxies for anxiety and depression, respectively) as nuisance variables. These covariates were selected based on prior literature (see Pliatsikas et al. (2018), Nikolin et al. (2021), and Lisica et al. (2022) for evidence of their relevance to the n-back task, and Van der Elst et al. (2006), Epp et al. (2012), and Menon et al. (2024) for their relevance to the Stroop task). Given the paucity of literature on the MST, the same covariates were assumed to be relevant for it as well. We avoided automatic selection techniques like stepwise regression due to their tendency to produce biased estimates and inflated p-values, among other issues, as described in Harrell (2015b). Interaction terms, though plausible, were not included due to the sample size being insufficient to test interaction effects reliably (Gelman, 2018).
ii. (ii) **Mental Health Metrics:** To assess associations of COVID-19 status and severity with our individual mental-health metrics and Mental Composite Score (MCS), we modeled the metrics as follows:

a. **PANAS PA and NA, GAD, and PHQ:** These metrics, being ordinal, were modeled using Ordered Logit Models (OLM) (i.e. Proportional Odds Logit Model). Additionally, GAD and PHQ scores, having a clinical cutoff at 3, were binarized and modeled using Logit Models (LM); to address separation issues, we applied mean bias reduction (Kosmidis et al., 2019) and obtained more stable estimates of the regression parameters.
b. **Mental Composite Score:** The metric, being continuous and unbounded, was modeled using a Multiple Linear Regression Model (MLRM). The same two-tier comparison strategy, i.e., (a) infected participants vs. healthy controls and (b) Mdrt+ vs. mild cases, with the same control variables, was applied as in the case of cognitive metrics. As before, we avoided automatic methods (Hosmer et al., 2013c) and relied on the clinical literature (Akhtar-Danesh and Landeen (2007); Kessler et al. (2001); Jayasankar et al. (2023)) to include age, gender, region, education, and presence of physical comorbidities as nuisance variables.

#### 2.3.3. Model Assumptions and Validation

i. **MLRM**: Multiple Linear Regression Model requires (a) independence of observations, (b) homogeneity of residual variance, and (c) normality of the pooled residuals, in order of decreasing priority (Zuur et al., 2009). Additionally, the model must exhibit acceptable goodness-of-fit. Independence is ensured by the structure of our dataset, where each subject contributes only one observation. For the remaining assumptions, we avoid blunt-edged statistical tests (Zuur et al., 2009) and instead rely on visual checks.

Except for *d^′^* (0-back), Q-Q plots of the residuals for any of the metrics in our models did not suggest any significant deviation from normality; simulation studies and practical experience across empirical studies attest that minor violations of the normality assumption, as seen in our examples, are allowed (Knief and Forstmeier, 2021). Likewise, plots of standardized residuals versus fitted values and scale-location plots showed no evidence of heteroskedasticity. We also verified that our models were not unduly influenced by outliers. To assess goodness-of-fit, we conducted F-tests comparing the fitted model with the intercept-only model; all models were significant at α = 0.05.

(ii) **BRM:** The Beta Regression Model requires that observations are independent and that the dependent variable is continuous and restricted to the open unit interval (0, 1) (Ferrari and Cribari-Neto, 2004). Independence of observations is, again, ensured by the study design. Model adequacy was assessed using the six standard diagnostic plots (Cribari-Neto and Zeileis, 2010). Across all cases, the Pearson residuals did not show any obvious pattern, with no evidence of misspecification (i.e., lack of GoF), heteroskedasticity beyond that accounted for by the precision parameter, or observations exerting disproportionate influence.
(iii) **OLM:** Ordered (ordinal) Logit Model requires that observations are independent and that the proportional odds assumption holds. Additionally, the model must demonstrate an acceptable goodness-of-fit. The first requirement continues to be met by the very nature of our data while the second is typically evaluated by Brant’s Wald test (Hosmer et al., 2013b). In our case, the test returned p-values less than 0.05 for all the models; however, given the very low expected frequencies in the contingency structure of our data (80% cells had values less than 0.01) due to sparse outcomes in many of our covariates, the statistical test could not be relied upon to evaluate our models (Conover, 1999).

When we plotted observed and model-estimated means of each predictor across outcome levels of the ordinal outcome, albeit in a simple univariate way, we found a total lack of monotonicity and overlap between the curves, indicating potentially severe violations of the proportional odds assumption (Harrell, 2015c). Indeed, the score-residual and partial-residual plots also indicated substantial departures from the expected patterns (Harrell, 2015c). Although such violations do not necessarily render an ordinal model wholly uninformative (Harrell, 2020), they imply that a single common odds ratio may not adequately summarize associations across all outcome thresholds. We therefore treat coefficient-level inference from these models, particularly isolated findings, as tentative.

(iv) **LM:** The Logit Model requires independence of observations and a linear relationship between the log odds of the dependent variable and each of the continuous covariates (Harrell, 2015a). Additionally, the model must demonstrate an acceptable goodness-of-fit. The first requirement continues to be met by the very nature of our data. The second requirement could not be resolved despite adding spline terms for our continuous variables (which improved the fit of our model only in a single case) and adding pairwise interactions (Harrell, 2015a).

Thus, instead of odds ratios, we only report the effect size (Average Marginal Effects; see Section 2.3.5 for details) as they are not only more robust to small sample size and nonlinear predictor functions (Bergtold et al., 2017) but also less sensitive to the sampled data and the model specification (Norton and Dowd (2017); but see Harrell (2021) for concerns on biases arising out of sample unrepresentativeness). We also verified that our models were not unduly influenced by outliers (Hosmer et al., 2013a). To evaluate goodness-of-fit (and diagnostic discrimination), rather than relying on the widely criticized Hosmer-Lemeshow test, we calculated the area under the Receiver Operating Curve (ROC) (Harrell, 2015a; Hosmer et al., 2013a) which exceeded 0.5 for all models, demonstrating better performance than chance.

For all diagnostic plots, see our code repository (for link, see Section 4).

#### 2.3.4. Bootstrapping and Confidence Interval

We sampled paired response–predictor observations with replacement 2000 times (Fox, 2015), used these resampled datasets to refit the model and obtain a distribution of regression coefficients, and used the bias-corrected and accelerated (BCa) bootstrap method (Efron, 1987) to construct our Confidence Interval (CI).

#### 2.3.5. Effect Size Indices

For continuous variables modeled using Multiple Linear Regression Models (MLRM), the regression coefficients themselves represent level-specific effect sizes and following Fox and Weisberg (2019), we do not standardize them. To quantify the contribution of each categorical variable as a whole, we report the partial omega squared (ω^2^), which estimates the expected proportion of population variance uniquely explained by that variable (Kroes and Finley, 2025; Richardson, 2011).

For categorical variables modeled using Ordinal Logit Models (OLM) or Logit Models (LM), we report the Average Marginal Effects (AME). AMEs represent differences in model-predicted outcomes between levels of a categorical predictor, averaging over a balanced grid of the remaining categorical predictors while holding numeric predictors at their means. Additionally, since for LMs, we report only the AME in lieu of the regression coefficients, we also report the corresponding p-values and confidence intervals using the BCa method.

For categorical variables modeled using Beta Regression Models (BRM), we do not report any effect size due to the lack of established metrics in the available literature.

#### 2.3.6. Reporting of Statistical Significance

While recognizing the limitations of exclusive reliance on p-values (Wasserstein and Lazar, 2016), we consider it to be a good yardstick in line with the latest statement by American Statistical Association (Benjamini et al., 2021) which suggests supplementing it with other measures. However, we acknowledge that the conventional 0.05 threshold is somewhat arbitrary (Amrhein and Greenland, 2017). Thus, while we treat results as significant only when the two-tailed p-value is below 0.05, we also report results with p-values near this threshold to highlight variables of interest for future research. Further, we supplement all our results with the confidence interval (CI). So, overall, we mention the following statistics when reporting any finding: (a) for GLMs, BRMs, and OLMs, unstandardized regression coefficient; for LMs, the Average Marginal Effect, (b) the corresponding two-tailed p-value, and (c) the corresponding bootstrapped 95% confidence interval (CI), rounded to two decimal places.

No formal correction for multiple testing was applied because the analyses concerned conceptually distinct outcomes (Bender and Lange, 2001) corresponding to separate hypotheses across working memory, pattern separation, attentional control, depression, anxiety, and affect. Although specific directional hypotheses were not preregistered, these tasks and measures were selected a priori based on prior work on COVID-19 and cognition, rather than as part of an exploratory screen across a broad range of outcomes.

#### 2.3.7. Directional Influence Sensitivity Analysis

Given the small Mdrt+ subgroup, we assessed whether statistically significant Mdrt+ effects were disproportionately driven by one or a few participants whose observations supported the estimated effect. For each outcome and contrast, we therefore conducted a sequential directional influence analysis using DFBETAS (Fox and Weisberg, 2019), which quantify the standardized change in a regression coefficient when an individual observation is omitted. At each iteration, we considered only Mdrt+ participants whose DFBETAS had the same sign as the original coefficient—that is, participants whose inclusion shifted the estimate in the observed direction—and removed the participant with the largest absolute DFBETAS. We then refitted the model and recalculated influence measures, repeating this procedure sequentially. We recorded the coefficient and two-sided *p*-value after each refit, stopping when *p* ≥ .10 or after five cumulative removals, and also noted the first iteration at which *p* ≥ .05. Thus, the procedure deliberately removed the Mdrt+ observations, providing the strongest support for each estimated effect, providing a stringent assessment of whether the result was driven by a small number of influential participants.

## 3. Results

Participants were distributed across a wide age range (*M* = 30.85 *±* 0.71; Fig. 2a). The groups were balanced with respect to gender and education levels (Table 2).

**Figure 2:**
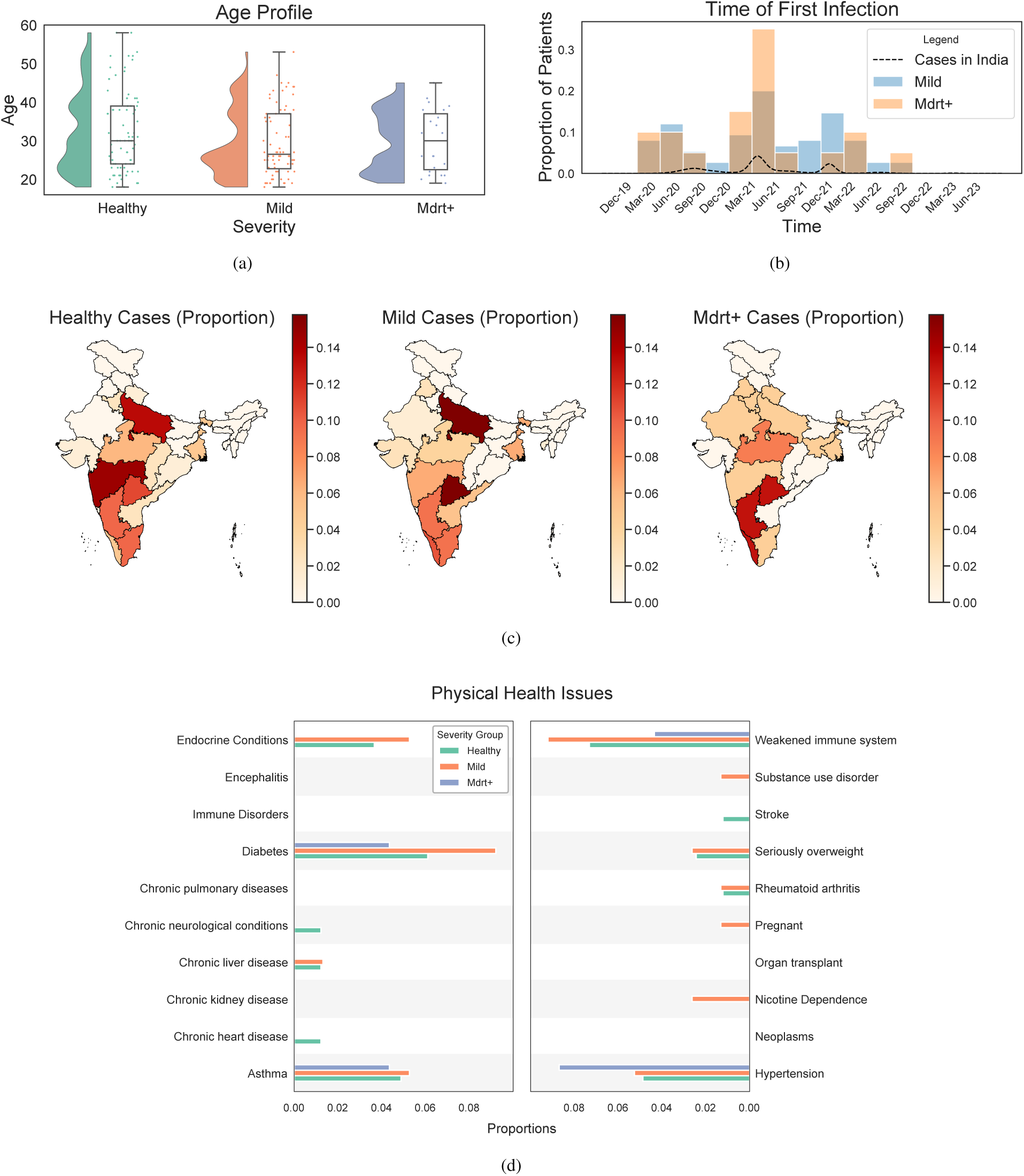
Demographic and clinical overview of the analytical cohort (*N* = 181), except panel **(b)**, which includes only infected participants with valid first-infection dates (*n* = 95). **(a)** Age distribution by COVID-19 severity. **(b)** Self-reported timing of first infection by COVID-19 severity, with a kernel density estimate (KDE) of pan-India COVID-19 cases obtained from WHO. **(c)** Geographic distribution by COVID-19 severity. **(d)** Prevalence of physical comorbidities by COVID-19 severity. The mirrored layout in panel **(d)** is used solely for visual compactness and does not represent a data transformation.

**Table 2:**
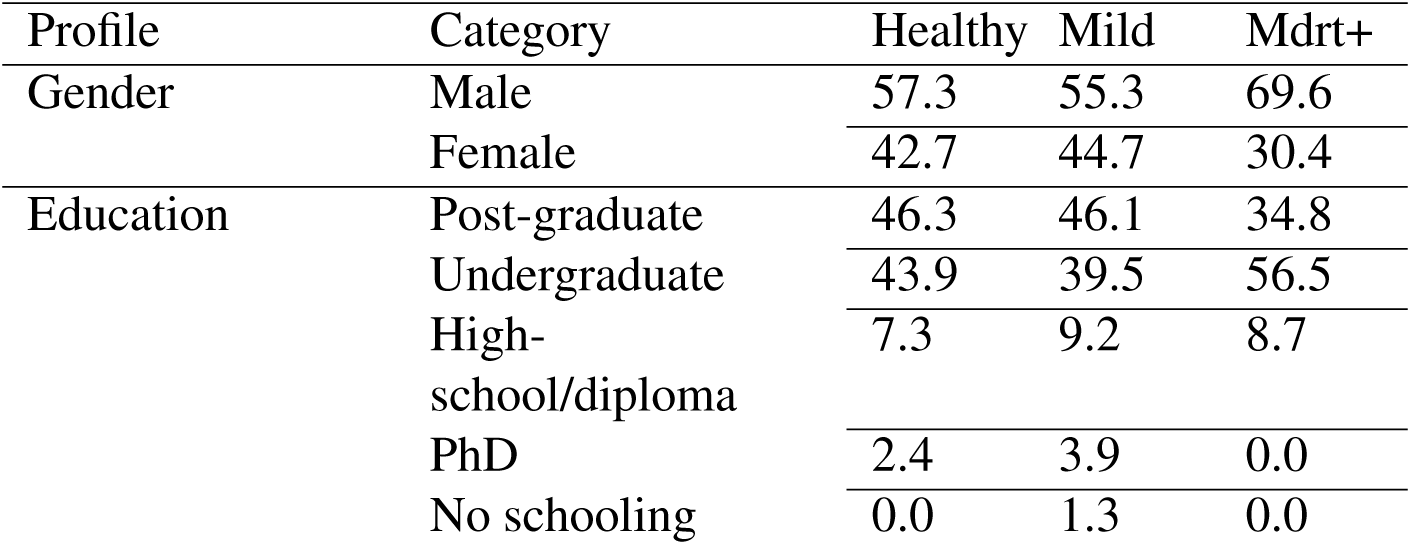
Demographic characteristics of the analytical cohort (*N* = 181), by COVID-19 severity group (%).

| Profile | Category | Healthy | Mild | Mdrt+ |
| --- | --- | --- | --- | --- |
| Gender | Male | 57.3 | 55.3 | 69.6 |
|  | Female | 42.7 | 44.7 | 30.4 |
| Education | Post-graduate | 46.3 | 46.1 | 34.8 |
|  | Undergraduate | 43.9 | 39.5 | 56.5 |
|  | High-school/diploma | 7.3 | 9.2 | 8.7 |
|  | PhD | 2.4 | 3.9 | 0.0 |
|  | No schooling | 0.0 | 1.3 | 0.0 |

The timing of first infection among our participants was broadly distributed across the COVID-19 pandemic, with peaks roughly coinciding with major infection surges in India, particularly in mid-2021 and early 2022 (Fig. 2b). This temporal correspondence not only supports the plausibility of the self-reported infection dates but also indicates that our cohort included infections acquired across all major national waves, and, in all likelihood, resulted from a mix of different SARS-CoV-2 variants.

Cases across all severity groups were distributed across India (except for the North and North-Eastern Cultural Zones (Ministry of Culture, Government of India, 2025)) indicating a spatially diverse cohort (Fig. 2c). Physical comorbidities, as self-reported, were rare across all the severity groups (Fig. 2d).

It is prudent to note that the vast majority of our infected participants would not have met the criteria for Long COVID, which requires symptoms persisting for at least three months after the initial infection and not attributable to alternative diagnoses (Greenhalgh et al., 2024). Most of our participants (87% of mild patients; 96% of mdrt+ patients) reported to have “fully recovered” at the time of the survey, with the preponderant majority (81% of mild patients; 74% of mdrt+ patients) claiming to have recovered within 2 months of being infected. Further, 17% of Mdrt+ participants and 50% of mildly infected participants had received at least one vaccination by the time of their first infection. This difference corresponded to a relative risk of 0.29 (Fisher’s exact test, *p* = .01), indicating that vaccination at first infection was associated with a 71% lower probability of Mdrt+ classification in our cohort and reinforcing the clinical consensus on the significant protective benefits of vaccination.

Overall, those who contracted COVID-19 of moderate-and-above severity fared the worst in all tasks. Participants performed well above chance across groups in all tasks (Table 3).

**Table 3:** Group mean cognitive-task scores and one-sided Wilcoxon signed-rank tests against chance-level performance in the analytical cohort (*N* = 181). **Note**: Task-specific sample sizes vary because not all participants completed every task.

| Severity | Count | $d'$ (0-back) | | $d'$ (1-back) | | $d'$ (2-back) | | LDI (MST) | | RT-IE% (Stroop) | |
| --- | --- | --- | --- | --- | --- | --- | --- | --- | --- | --- | --- |
| | | Score | $p$ | Score | $p$ | Score | $p$ | Score | $p$ | Score | $p$ |
| **Healthy** | 82 | 3.49 | < 0.005 | 2.11 | < 0.005 | 1.78 | < 0.005 | 0.15 | < 0.005 | 21.70 | < 0.005 |
| **Mild** | 76 | 3.53 | < 0.005 | 2.16 | < 0.005 | 1.83 | < 0.005 | 0.19 | < 0.005 | 20.42 | < 0.005 |
| **Mdr+** | 23 | 3.07 | < 0.005 | 1.00 | 0.01 | 0.54 | 0.01 | 0.10 | < 0.005 | 19.70 | < 0.005 |

A summary of the principal regression contrasts is provided in Table 4. In the following sections, we first report associations of infection severity with overall cognitive performance and mental health in the full analytical cohort, as measured using the composite scores. We then report corresponding associations with individual cognitive tasks and questionnaires when they were statistically significant. Finally, we examine the association between vaccination status at first infection and cognitive performance.

**Table 4:**
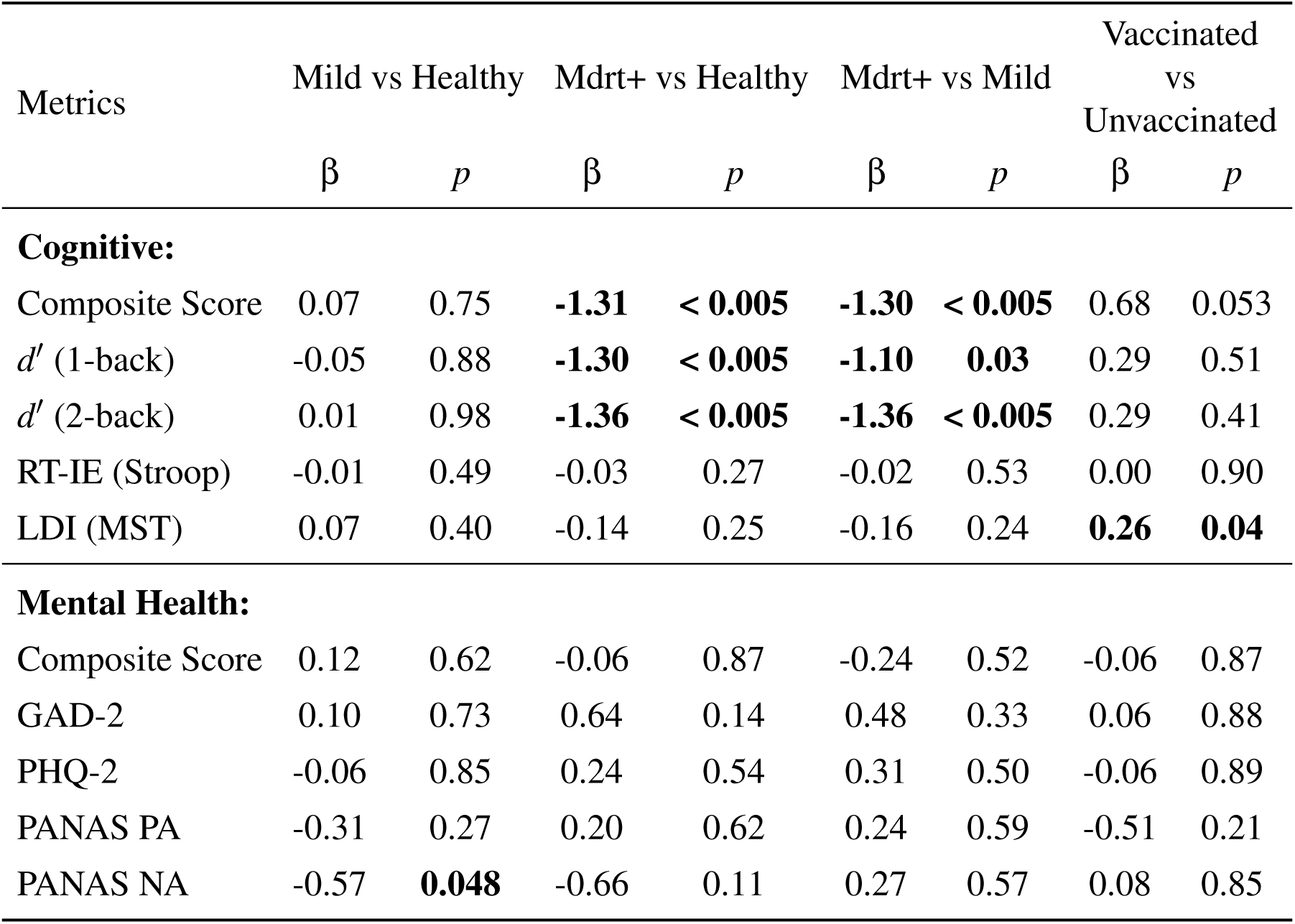
Regression results comparing cognitive and mental health metrics across COVID-19 groups. Significant results (*p <* 0.05) are boldfaced. The regression β comes from different models; for LDI, the coefficient of the transformed variable ((*LDI* + 1)*/*2) is reported.

### 3.1. Cognitive Performance

Patients who experienced COVID-19 of moderate and above severity (Mdrt+) demonstrated poorer global cognition compared to both healthy controls (β = *−*1.31; *p <* 0.005; 95% C.I. = [*−*1.94*, −*0.60]; ω^2^_p_) and mildly infected patients (β = *−*1.30; *p <* 0.005; 95% C.I. = [*−*2.08*, −*0.42]; ω^2^_p_), as shown in Fig. 3a. In contrast, mildly infected participants did not differ significantly from healthy controls (β = 0.07; *p* = 0.75; 95%C.I. = [*−*0.38, 0.52]). These results indicate that greater reported COVID-19 severity was associated with lower overall cognitive performance; the corresponding effect sizes were of moderate magnitude (Field, 2019) and indicate that severity differences accounted for a meaningful and reliable portion of the variance in cognitive performance.

**Figure 3:**
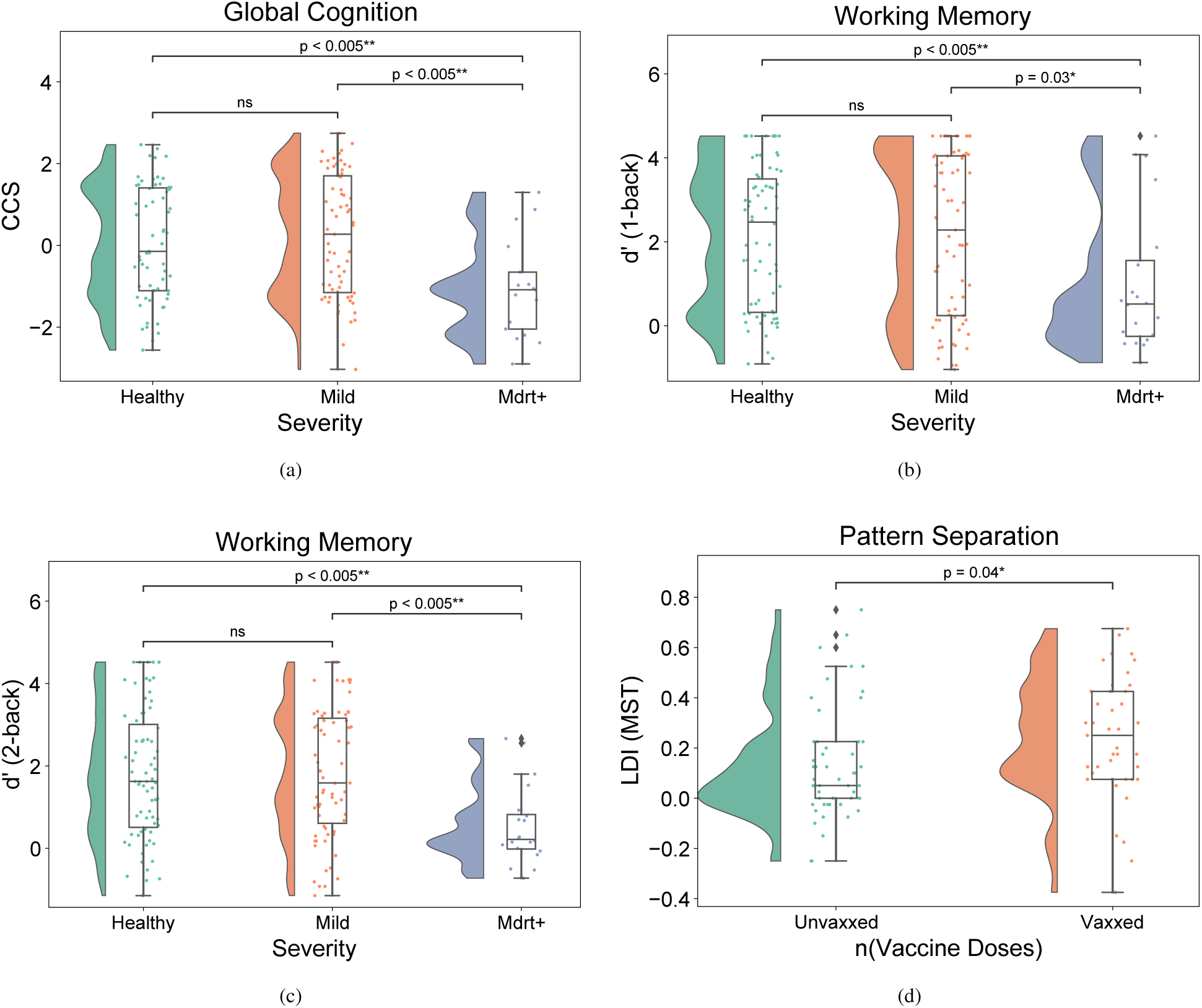
Differences in cognitive performance across COVID-19 severity and vaccination status. All *p*-values are from our regression models. **(a)** Cognitive Composite Scores were significantly lower in the Mdrt+ group than in healthy controls and mildly infected participants. **(b)** Performance on the 1-back task was significantly lower in the Mdrt+ group than in healthy controls and mildly infected participants. **(c)** Performance on the 2-back task was significantly lower in the Mdrt+ group than in healthy controls and mildly infected participants. **(d)** Participants vaccinated at the time of infection showed better MST performance, as indexed by the Lure Discrimination Index, than unvaccinated participants.

To identify the cognitive domains underlying the composite-score differences, we fitted regressions to the individual tasks. The lower composite scores in the Mdrt+ group were explained by lower performance on the 1-back task (Mdrt+ vs Healthy: β = *−*1.30; *p <* 0.005; 95% C.I. = [*−*2.13*, −*0.33]; ω^2^_p_ and Mdrt+ vs Mild: β = *−*1.10; *p* = 0.03; 95% C.I. = [*−*2.15, 0.12]; ω^2^_p_) and on the 2-back task (Mdrt+ vs Healthy: β = *−*1.36; *p <* 0.005; 95% C.I. = [*−*1.97*, −*0.77]; ω^2^_p_ and Mdrt+ vs Mild: β = *−*1.36; *p <* 0.005; 95% C.I. = [*−*2.08*, −*0.63]; ω^2^_p_), as shown in Fig. 3b and Fig. 3c, respectively. Mean response times did not differ significantly across the three groups for either task, suggesting that the between-group score differences did not reflect a speed–accuracy trade-off.

Thus, taken together, the observed cognitive decline in the Mdrt+ group can be primarily explained by a decline in working memory. To preclude the possibility that these findings depended on a small number of influential Mdrt+ participants (given our small group size), we conducted the sequential directional influence analysis as described in Section 2.3.7, cumulatively removing participants who were the most influential in the regression. All six Mdrt+ coefficients retained their negative direction, although their inferential stability varied. The Mdrt+ vs. mild 1-back contrast ceased to meet the conventional *p < .*05 threshold after one removal, whereas the remaining contrasts required three to five removals to cross this threshold or remained below it even after five removals. Precise details about inferential robustness with each successive removal can be seen in Appendix C.

### 3.2. Mental Health

Patients who contracted COVID-19 of a moderate and above severity did not show a statistically significant difference in overall mental health when compared to healthy controls (Mdrt+ vs Healthy: β = *−*0.06; *p* = 0.87; 95% C.I. = [*−*0.69, 0.54]; ω^2^_p_) or mildly infected patients (Mdrt+ vs Mild: β = *−*0.24; *p* = 0.52; 95% C.I. = [*−*0.98, 0.52]; ω^2^_p_). Expectedly, mildly infected patients did not show any difference when compared to the healthy controls (β = 0.12; *p* = 0.62; 95% C.I. = [*−*0.36, 0.60]). To assess whether COVID-19 was associated with specific aspects of mental health, we fitted regressions to the individual PHQ, GAD, and PANAS scores. No group contrasts were statistically significant except that mildly infected participants reported lower PANAS negative affect than healthy controls (β = *−*0.57, *p* = 0.048), contrary to the anticipated direction; however, this finding should be interpreted cautiously because the ordinal model exhibited substantial violations of the proportional-odds assumption and sparse outcome levels (Section 2.3.3).

To ensure that the presence of physical comorbidities was not masking potential mental health effects, we re-ran our regression models for both the mental composite score and individual questionnaires without controlling for comorbidities. This step addressed the possibility that comorbidities acted as mediators—lying on the causal pathway from disease severity to mental health outcomes (for instance, COVID-19 increasing the risk of physical illness, which in turn affects mental health)—rather than as independent confounders. The results remained unchanged after omitting physical comorbidities: the mild-versus-healthy difference in PANAS negative affect persisted, while all other group contrasts remained nonsignificant. Nevertheless, this isolated finding remains subject to the same limitations, as described before.

However, when modeling GAD as a binary outcome using the clinical threshold (score *≥* 3), we found statistically significant elevation in the likelihood of anxiety among patients with moderate and above severity compared to healthy controls (Mdrt+ vs Healthy: AME = 0.25; *p* = 0.02; 95%C.I. = [0.03, 0.46]). The difference in results from the OLM, most likely arose due to the sparsity across individual response levels (which can substantially affect statistical power) and/or the extensive violations of the proportional odds assumption. Nonetheless, given that the assumptions of the Logistic model were violated too (Section 2.3.3), we suggest extreme caution in interpreting these findings.

### 3.3. Association of vaccination with cognitive performance

Vaccination status as of first infection was treated as a binary variable (unvaccinated vs vaccinated), with all participants who had received at least one dose grouped together because the numbers in the individual strata were too sparse for reliable standalone contrasts. Vaccinated participants had higher estimated global-cognition scores than unvaccinated participants, although this difference did not meet the conventional significance threshold (Vaccinated vs None: β = 0.68; *p* = 0.053; 95% C.I. = [*−*0.06, 1.32]; ω^2^_p_). This difference could be explained by their superior performance in the MST Task over unvaccinated counterparts (β = 0.26; *p* = 0.04; 95% C.I. = [*−*0.01, 0.46]), as shown in Fig. 3d. The improvement was not due to a speed-accuracy tradeoff; there were no significant differences between the average time taken by patients across the two groups. Thus, vaccination status at first infection was associated with better pattern-separation performance.

Owing to the skewed distribution of vaccination status across severity groups (most Mdrt+ participants were unvaccinated), we examined whether the estimated association with vaccination was sensitive to this imbalance in infection severity, notwithstanding the inclusion of severity as a covariate, by repeating the analysis in a sub-cohort comprising only mildly infected participants. The estimate in the mild group narrowly missed the conventional threshold for statistical significance (*p* = 0.051), but was consistent with the association observed in the full sample, suggesting that the finding was not driven by the imbalance in severity. Furthermore, to assess the sensitivity of our findings to model specification, we repeated the analyses in both samples using a Multiple Linear Regression Model (MLRM) instead of a Beta Regression Model (BRM). The MLRM yielded slightly stronger statistical evidence, with significant vaccination associations in both the full infected sample and the mild-only sub-cohort (see Appendix D).

## 4. Discussion

Situated within the South Asian context, our findings add to the scarce literature (see Chakrabarty et al. (2024) and Gopinath et al. (2025) for exceptions) on the long-term cognitive performance of survivors of COVID-19. In line with existing evidence from other regions (Cui et al., 2023; Aretouli et al., 2025), we observe that individuals who experienced moderate-to-severe symptoms of COVID-19 exhibit persistent deficits in working memory—as assessed by the 1-back and 2-back tasks—well beyond the acute phase and extending at least six months post-infection. Notably, these impairments were absent among mildly infected individuals, suggesting that the long-term outcomes of COVID-19 are primarily associated with illness severity. Importantly, time since infection was not significantly associated with working-memory performance, indicating that the observed deficits may be persistent rather than progressively improving, consistent with findings from the latest meta-analysis on long-COVID (Hou et al., 2025) but contrary to the trends reported in (Cui et al., 2023). Further, we observed no significant cognitive slowing (see Zhao et al. (2024) and Weinerova et al. (2025) for mixed evidence) in either infected group relative to healthy controls on any task.

However, unlike multiple studies that document impaired selective attention of patients with a history of COVID-19 upon administration of the Stroop Task (see Panagea et al. (2024) and Table 3 in Nasir et al. (2025) for an overview), we observed no corresponding between-group difference. Likewise, although studies employing the Mnemonic Similarity Task (MST) in COVID-19 populations are extremely rare, our findings diverge from Meyer and Zaiser (2025), who reported significant hippocampal deficits in COVID-19 patients compared to healthy controls. These differences, we believe, are unlikely to stem from minor variations in analytical approach or demographic composition but instead primarily reflect two features of our study design: (a) our broader criteria for classifying disease severity, whereas many previous studies define severe disease largely or exclusively by ICU admission, thereby selecting a more severely affected group in whom cognitive deficits may be easier to detect; and (b) our participant recruitment strategy, which differed from that of most previous studies, many of which tested participants within six months of infection or specifically recruited individuals with long COVID. Notably, among the Stroop studies summarized in Nasir et al. (2025), Bungenberg et al. (2022) employed the recruitment strategy most comparable to ours and, like us, found no corresponding impairment. We elaborate on both the aspects below.

Much of the existing literature on COVID-19-related cognitive dysfunction (see Voruz et al. 2022, Vakani et al. 2023, de Pádua Serafim et al. 2024, Wood et al. 2024, and Demmer et al. 2025 for individual studies; Austin et al. 2024 and Aretouli et al. 2025 for meta-analyses) classifies disease severity according to the latest WHO guidelines (WHO Working Group on the Clinical Characterisation and Management of COVID-19 infection, 2020), wherein the moderate and severe categories are operationalized based on hospitalization status. Correspondingly, these studies report more significant cognitive impairments across multiple domains among hospitalized patients compared to non-hospitalized ones. Our study diverges in an important methodological respect: we adopted an older classification schema that assessed severity without wholly relying on hospitalization (*≈* 87% of our moderate-to-severe cases were managed outside of hospital settings). This enabled us to observe substantial working memory impairments in patients who had not been hospitalized yet had otherwise borne the brunt of the infection, while at the same time possibly explaining why we did not observe any significant impairment in selective attention or hippocampal integrity within our cohort.

Our findings thereby reveal an important blind spot in current research paradigms and prompt further reflection. During the height of COVID-19 waves in India and other low-resource settings, hospitalization was not always feasible even for severe cases due to healthcare system overload (Zargar, 2021; Arora, Neha and Ravikumar, Sachin, 2021). Relying exclusively on hospitalization as a marker of severity can therefore systematically exclude a vulnerable segment of the population who experienced the full severity of illness without access to hospital-based care. Even beyond the constraints of such contexts, select prior studies report cognitive impairments in non-hospitalized individuals: He et al. (2023) reported cognitive deterioration even among non-hospitalized patients with clinically moderate or severe COVID-19; Kirchberger et al. (2023) found similar impairments in patients with a mild disease profile, while Miskowiak et al. (2023) found no significant difference in the extent of impairment between hospitalized and non-hospitalized patients approximately seven months after infection. Our findings align with the thrust of these results, and the effect sizes of global cognitive impairment we observe for the moderate-to-severe group are comparable to those reported for hospitalized samples in studies such as Ollila et al. (2022). Collectively, these results caution against using hospitalization as an automatic proxy for illness severity in studies of long-term cognitive sequelae, especially in low-resource regions with poor public health infrastructure.

Another methodological distinction lay in how participants were recruited. Most studies that focus on the long-term effects of COVID-19 (e.g., Herrera et al. 2023, Hampshire et al. 2024, Boutet et al. 2025; particularly see Recommendation 1 in Becker et al. (2023) on selection bias) recruit individuals who self-report cognitive complaints in tune with the definition of long-COVID. Such studies, if used to draw conclusions about the long-term impact of COVID-19 in the broader population, can not only inflate the extent of impairments but also obscure subtler dysfunctions. Further, our findings, alongside those of Ariza et al. (2023); Meyer and Zaiser (2025); Voruz et al. (2022), suggest that individuals who consider themselves fully recovered (to reiterate, 96% of Mdrt+ patients reported to have “fully recovered” at the time of the survey) can show measurable deficits in cognitive domains; this points to a significant disconnect between subjective recovery and true cognitive functioning and suggests that self-perception may not be a reliable indicator when screening for long-COVID.

Although MST performance did not differ significantly between infected participants and healthy controls, vaccination status at the time of infection was associated with better MST performance six months or more post-infection, suggesting a potential protective effect on hippocampal-dependent pattern separation. At first glance, this may appear counterintuitive, i.e., if MST performance is not measurably reduced as a function of COVID-19 severity in our sample, why would vaccination confer any protection? One possibility is that the post-COVID group contains a mix of people: some more vulnerable to hippocampal disruption and others less so, regardless of clinical severity. If vaccination protected the more vulnerable subgroup, then a benefit would emerge only when comparing vaccinated with unvaccinated individuals, even if mean MST performance by severity alone appears unaffected. This interpretation is speculative, and given the limited literature addressing cognitive protection conferred by COVID-19 vaccines (Zhao et al., 2023), we urge caution. Nonetheless, our finding is broadly consistent with a recent proposal, based on indirect evidence from animal studies of influenza and BCG vaccines, that COVID-19 vaccination may preserve or enhance adult hippocampal neurogenesis (Kumar et al., 2023; Goldstein, 2023), a mechanism that could support improved pattern separation. These converging observations, while preliminary, point to a potentially important and currently underexplored avenue for future research on long-term neurocognitive outcomes following COVID-19.

Although our results provide important initial insights, several limitations must be acknowledged. First, the small sample size limited our ability to examine plausible interaction effects and may also have obscured subtler cognitive impairments among mildly infected participants. Second, because the study was cross-sectional, pre-infection cognitive baselines were unavailable; thus, despite adjustment for a broad set of measured confounders, residual confounding by unmeasured factors remains possible and the observed differences cannot be attributed conclusively to COVID-19. Third, our clinically defined severity groups reduced dependence on hospitalization, which was an unreliable proxy during periods of hospital overload, but relied on retrospective self-report rather than clinical adjudication and may therefore have introduced recall and classification error. Fourth, although the N-back, Stroop, and MST index important cognitive domains, they do not capture the full range of difficulties reported in post-COVID populations, including verbal fluency, planning, and processing speed. Fifth, although our finding of no enduring affective or mood disturbances across groups is consistent with reports that cognitive and psychiatric sequelae can be dissociable (but see Rudenstine et al. (2022) and Vallée et al. (2025)), violations of the assumptions underlying our mental-health models preclude firm conclusions. Sixth, the absence of direct supervision during remote testing may have introduced variability in adherence to task instructions and participant engagement. Lastly, our online recruitment strategy likely favored younger, more educated, and technologically proficient participants; the findings should therefore not be generalized to the Indian population at large.

Future research, especially in the subcontinent, should prioritize retrospective or cohort-based longitudinal approaches, leveraging existing health records where possible to reconstruct pre- and post-infection trajectories; studies incorporating repeated follow-ups in previously infected cohorts (Liu et al., 2022; Taquet et al., 2024; Liu et al., 2024) appear to be valuable in tracking recovery trajectories and identifying delayed-onset impairments. Broader cognitive assessments are also needed, using more comprehensive test batteries and, optimally, incorporating EEG and/or fMRI measures. Sampling strategies should actively include non-hospitalized populations, who are often underrepresented in clinic- and hospital-based studies, and should aim for sample sizes large enough to permit fine-grained comparisons across viral strains (Latifi and Flegr, 2024; Lynch et al., 2026). Further, larger cohorts with more complete vaccination histories would allow future studies to examine whether vaccine type, dose number, and the timing of vaccination relative to both infection and cognitive assessment modify the observed associations in our study.

Overall, our results challenge the growing public and institutional tendency to forget the COVID-19 pandemic. While acute symptoms may fade and many recover without complications, our findings reinforce that moderate-to-severe COVID-19 can have chronic cognitive consequences that persist well beyond subjective assessments of recovery. This underscores the importance of sustained vigilance, cognitive screening, and possible rehabilitation support (see Knopman et al. (2026) for a failed attempt) even in a post-pandemic world. Disregarding these risks could leave a substantial portion of the population under-served and unrecognized in their cognitive struggles, with potentially far-reaching consequences for individual productivity and national economy.

## Contributions

**Aruneek Biswas**: Writing – original draft; Formal Analysis (lead); Data Curation; Visualization **Vivek Pamnani**: Investigation; Formal Analysis (supporting); Software; Writing – Review & Editing (supporting) **Priyanka Srivastava**: Conceptualization; Funding Acquisition; Supervision; Project Administration; Writing – Review & Editing (supporting) **Vishnu Sreekumar**: Conceptualization; Funding Acquisition; Supervision; Project Administration; Writing – Review & Editing (lead)

## Data Availability

Anonymized raw data and code will be provided to reviewers and released upon publication.

## Acknowledgements

We would like to thank Pritha Ghosh and Sriya Ravula for providing useful feedback on earlier versions of the manuscript. We acknowledge IHub-Data, IIIT Hyderabad (IIIT-H/IHub/Project/ Healthcare/2021-22/H2-004; PS and VS) and IIIT Hyderabad Faculty Seed Fund (IIIT/R&D Office/ Seed-Grant/2021-22/018; VS) for supporting this work.

## Conflict of Interest

None of the authors have any COI to report.

## Appendix A. Survey Questionnaire

### Appendix A.1. Questions on Demographics

- To which gender identity do you mostly identify?

– Man
– Woman
– Transgender Woman
– Transgender Man
– Non-Binary Gender
– Prefer not to say
– Prefer to self-describe:
- How old are you?

– (Allowed range: 18–70)
- What kind of area do you live in?

– City
– Sub-urban
– Rural
- **Choose the state/union territory where you are residing currently:**

– One had to pick from a list of Indian states and UTs.
- **Choose the option that best describes your employment status:**

– Working (full time)
– Working (part-time)
– Not working (on a leave)
– Not working (looking for work)
– Not working (retired)
– Not working (student)
– Not working (other)
– Volunteer workers
– Casual and temporary employees
– Home maker
– Home-keeping
– Trainees/apprentices
– Employee on probation
– Foreign Employees
– Contract / Subcontract workers
- **Select the option which best describes your latest education status:**

– No Formal School Education
– Vocational Education or Skill Training (e.g., Carpentry, Mechanic skill learning)
– Level IV/V or lower
– Level VI – Level X
– Level XI – Level XII or Higher Secondary
– Graduation
– Post Graduation
– Diploma
– PhD
– Professional Courses (e.g., MBA)

### Appendix A.2. Select Questions on Infection, Recovery, and Vaccination

(Questions in red were displayed if the answer to the question on having symptoms of COVID-19 was answered in affirmative):

- **Please select one option that indicates your current COVID-19 vaccination status:**

– Not Vaccinated
– Partially Vaccinated (one dose only)
– Fully Vaccinated
– Fully Vaccinated with one booster dose
– Fully Vaccinated with two booster doses
- Have you ever been tested positive for COVID-19?

– Yes (when tested using swab/RTPCR/Scan/antigen/others)
– No (when tested using swab/RTPCR/Scan/Antigen/others)
– Awaiting results (when tested using swab/RTPCR/Scan/Antigen/others)
– Did not get tested
- **Have you had or suspect you have had symptoms of COVID-19?:**

– Yes
– No
- **When did you experience COVID-19 infection or symptoms?** (If more than once, pick the 2–3 most recent instances.)

– Date-time picker
– Date-time picker
– Date-time picker
- Please select one of the following according to the severity of your COVID-19 symptoms:

**– Asymptomatic or Presymptomatic Infection**: Individuals who test positive for COVID-19 but who have no symptoms that are consistent with COVID-19.
**– Mild Illness**: Individuals who have any of the various signs and symptoms of COVID-19 (e.g., fever, cough, sore throat, malaise (a general feeling of discomfort, illness, or uneasiness whose exact cause is difficult to identify), headache, muscle pain, nausea, vomiting, diarrhea, loss of taste and smell) but who do not have shortness of breath, dyspnea, or abnormal chest imaging.
**– Moderate Illness**: Individuals who show evidence of lower respiratory disease during clinical assessment or imaging and who have an oxygen saturation (SpO2) *≥* 94% on room air at sea level.
**– Severe Illness**: Individuals who have SpO2 < 94% on room air at sea level, a ratio of arterial partial pressure of oxygen to fraction of inspired oxygen (PaO2/FiO2) < 300 mm Hg, a respiratory rate > 30 breaths/min, or lung infiltrates > 50%.
**– Critical Illness**: Individuals who have respiratory failure, septic shock, or multiple organ dysfunction.
- Please select the appropriate medical course of action(s) that you have experienced. Select one or more of the following:

– Home Quarantine/Isolation
– Home Stay with Medical Care/Support
– Hospitalization:

_*_ Hospitalization (General Ward/Special Ward)
_*_ Hospitalization with Intensive Care Unit (ICU) / Intensive Treatment Unit (ITU) / Critical Care Unit (CCU) / Intensive Coronary Care Unit (ICCU)
– Mechanical Support:

_*_ Continuous Oxygen supplement
_*_ Oxygen Support but no continuous requirement
_*_ Ventilator support / Invasive mechanical ventilator support
_*_ High-flow nasal support
- Do you feel fully recovered?:

– Yes
– No
– Not sure
- Please mention the duration of post-COVID symptoms (e.g., fatigue, loss of taste and smell, etc.) you experienced since the infection onset. Choose one which represents the duration of recovery most closely.:

– I am still experiencing post-COVID symptoms
– Less than 1 week
– 1–2 weeks
– 3–4 weeks
– Almost 2 months
– Almost 3 months
– Almost 4 months
– Almost 5 months
– Almost 6 months
– More than 6 months
– More than 12 months
– More than 18 months
– More than 24 months
- When did you experience the COVID-19 symptoms?

– Before vaccination
– After partial vaccination (one dose only)
– After full vaccination (two doses)
– After full vaccination with one booster dose
– After full vaccination with two booster doses
- Which vaccine did you take?

– Covishield
– Covaxin
– Sputnik-V
– Not listed? Enter the name here:
- **Please enter the month and year of your first dose of vaccination to the best of your memory:**

–
- **If you could not remember the month/year of your first dose of vaccination and skipped the question, consider visiting the official CoWIN portal** ( https://selfregistration.cowin.gov.in) **to check your vaccination history.**:

–

## Appendix B. Exclusion criterion

To ensure data quality, we imposed the following filters, successively:

1. **Inattentive and Non-serious Participants**: Seven participants were excluded for failing two or more of the eleven cognitive and attention-related checks. These checks included outlying response times (z-score *≥* 2 for the five tasks), task performance outliers (IQR for the five task metrics), and incorrect response to an attention-check question.
2. **Inconsistent COVID-19 History**: Sixteen participants were excluded for self-reporting a positive COVID-19 test while also indicating that they had never suspected having COVID-19.
3. **Invalid Vaccination Year**: Seven participants were excluded for reporting a spurious vaccination year, even after being asked to cross-reference with the CoWin database.
4. **Contradictory Vaccination and Infection Timeline**: Two participants were excluded for reporting a vaccination date that preceded their infection but later claiming to have experienced COVID-19 symptoms before vaccination.
5. **Inconsistent COVID-19 Severity and Treatment Reports**: Seven participants were excluded for reporting mild or moderate COVID-19 severity but indicating treatment levels inconsistent with their severity classification (e.g., hospitalization or ventilator use for mild cases).
6. **Self-reported Mental-health Diagnosis**: Sixteen participants were excluded after self-reporting a mental-health condition that was likely to substantially affect performance on the cognitive battery (e.g., dementia, ADHD, OCD, bipolar disorder, or depression). These reports were not independently verified. Although these participants performed approximately at chance level only on the 1-back task (median *d^′^* = 0; one-sided Wilcoxon signed-rank *p* = .08), they were excluded according to the prespecified eligibility criterion.

## Appendix C. Results of the Directional Influence Sensitivity Analysis

The sequential directional influence analysis described in Section 2.3.7 produced the deletion paths reported in Table C.1:

**Table C.1:**
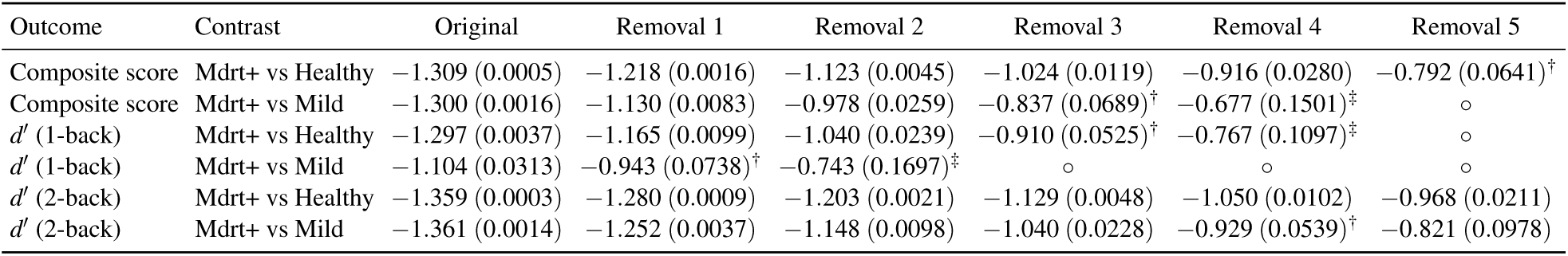
Sequential directional DFBETAS sensitivity analysis of the significant Mdrt+ coefficients in Table 4. Entries are unstandardized regression coefficients, with two-sided *p*-values in parentheses. ^†^ denotes the first value reaching *p* ≥ .05; ^‡^ denotes the first value reaching *p* ≥ .10; *◦* denotes that the analysis had already stopped at some preceding removal after reaching *p* ≥ .10.

## Appendix D. Robustness Check: Benefit of vaccination for LDI performance

**Table D.2:** Note that the regression betas are not comparable across the two model types; for Beta Regression Models (BRMs), the coefficient for the transformed outcome, (*LDI* + 1)*/*2, is reported.

| Model | Included Patients | Beta | P-value | CI Lower | CI Upper |  |
| --- | --- | --- | --- | --- | --- | --- |
| BRM | (Mdr<sup>+</sup>, Mild) | 0.26 | **0.04** | -0.01 | 0.46 | ← Primary specification |
| BRM | Mild | 0.30 | 0.051 | -0.03 | 0.54 |  |
| MLRM | (Mdr<sup>+</sup>, Mild) | 0.13 | **0.03** | 0.01 | 0.22 |  |
| MLRM | Mild | 0.15 | **0.04** | -0.01 | 0.26 |  |

## Footnotes

2 Can be accessed at https://www.cloudresearch.com as of 13 November 2025.

3 We did not collect sex assigned at birth. However, sex- and gender-related factors may differentially affect cognitive outcomes, with interactions between the two also reported (Cartier et al., 2024).

## Notes

### Competing Interest Statement

The authors have declared no competing interest.

### Author Declarations

IIIT Hyderabad Research Ethics Committee approved our work.

### Summary of Updates

This revision clarifies recruitment strategies, group construction, severity classification, questionnaire scoring, statistical modelling, and interpretation, and, in particular, expands the study limitations and sensitivity analyses. We also corrected inconsistencies in terminology, reporting, figures, and tables and removed causal language throughout in light of our design. The reported full-cohort sample is also corrected from N = 177 to N = 181 by retaining four previously excluded participants in analyses not requiring reliable infection dates; no study conclusions changed.

